# Health Technology Assessment of Diagnostic Tests: A state of the art review of methods guidance from international organisations

**DOI:** 10.1101/2022.05.17.22275215

**Authors:** Lavinia Ferrante di Ruffano, Isobel M Harris, Zhivko Zhelev, Clare Davenport, Sue Mallett, Jamie Peters, Yemisi Takwoingi, Jon Deeks, Chris Hyde

## Abstract

**Objectives:** To identify which international HTA agencies are undertaking evaluations of medical tests, summarise commonalities and differences in methodological approach, and highlight examples of best practice.

**Methods:** A methodological review incorporating: systematic identification of HTA guidance documents mentioning evaluation of tests; identification of key contributing organisations and abstraction of approaches to all essential HTA steps; summary of similarities and differences between organisations; and identification of important emergent themes which define the current state of the art and frontiers where further development is needed.

**Results:** Seven key organisations were identified from 216 screened. The main themes were: elucidation of claims of test benefits; attitude to direct and indirect evidence of clinical effectiveness (including evidence linkage); searching; quality assessment; and health economic evaluation. With the exception of dealing with test accuracy data, approaches were largely based on general approaches to HTA with few test-specific modifications. Elucidation of test claims and attitude to direct and indirect evidence are where we identified the biggest dissimilarities in approach.

**Conclusions:** There is consensus on some aspects of HTA of tests, such as dealing with test accuracy, and examples of good practice which HTA organisations new to test evaluation can emulate. The focus on test accuracy contrasts with universal acknowledgement that it is not a sufficient evidence base for test evaluation. There are frontiers where methodological development is urgently required, notably integrating direct and indirect evidence and standardising approaches to evidence linkage.

## Introduction

Health Technology Assessment (HTA) of tests differs to therapeutic interventions. One of the most important differences is that medical testing rarely improves health outcomes directly. Testing is usually part of a complex clinical pathway where test results guide treatment decisions, which include a variety of medical actions and processes. Medical tests have the potential to improve patient outcomes if improvements in accuracy (reductions in false positives and false negatives) are translated into more appropriate diagnoses (diagnostic yield) and more appropriate treatment (therapeutic yield). Treatment effectiveness ultimately determines the degree to which improvements in therapeutic yield will result in improved patient outcomes [1]. Medical tests may also improve patient outcomes by mechanisms other than accuracy. For example, tests may offer similar accuracy at reduced cost, simplify healthcare delivery thereby improving access, improve diagnostic confidence and improve diagnostic yield, reduce time to diagnosis, lower anxiety, reduce uncertainty or improve safety [2].

Assessing test effectiveness therefore requires evaluation and comparison of complex test-treatment management strategies [3-5]. Direct (end-to-end) evidence of the effect of tests on patient outcomes as might be captured by a randomised controlled trial (RCT) is scarce [6]. In the absence of RCT evidence a ‘linked evidence approach’ methodology has been advocated [7] to evaluate the clinical utility of medical tests: using decision analytical modelling to integrate evidence [8] on each component of a test–treatment pathway.

The number of international HTA organisations with a remit including non-pharmacologic technologies has increased over the last decade [9-11]. Concurrently, methodological research has defined the types of research required to evaluate medical tests [12,13] and methods to undertake HTAs of tests have been developed. These include, the launch of the UK’s National Institute for Health and Care Excellence Diagnostics Assessment Programme (NICE DAP) in 2010 [14], and a programme of methods development for conducting HTAs of tests by the USA’s Agency for Healthcare Research and Quality (AHRQ) [15].

Existing reviews of international HTA methods have demonstrated that many organisations rely on established HTA systems set up for evaluation of pharmaceuticals and apply these to tests with little or no modification [10,11,16]. Substantial variation in evidence requirements for HTA of tests across international organisations has also been noted [11,17]. Reviews of UK National Institute of Health Research HTA reports [17,18] have demonstrated that, whilst uptake of appropriate meta-analytic methods for test accuracy evidence has improved over the last decade, the extent to which these are incorporated in economic models remains variable, as do methods for ascertaining and using non–test accuracy evidence. No improvement was noted in consideration of the effects of threshold and dependency when evaluating test combinations [17].

We are not aware of any reviews that have sought to summarise the methods used by HTA organisations to perform HTAs of tests. Mapping the extent, features and variation in approach to HTA of tests is key to identifying and sharing best practice, preventing duplication of effort and to informing an international research agenda.

Drawing on methods already used to identify and map international HTA activity for pharmaceuticals and medical devices [10,16] the aims of this review were to identify which international HTA agencies are undertaking evaluations of medical tests, summarise commonalities and differences in methodological approach, highlight examples of best practice as well as areas requiring methodological development.

## Methods

We performed a descriptive analysis of publicly available guidance documents of international HTA organisations who evaluate medical tests. For organisations providing detailed test–specific methods guidance this consisted of a thorough examination of methods and process documents. This was complemented by a shorter systematic interrogation of methods guides for all other organisations to identify additional innovative methods for HTAs of tests.

### Identifying HTA organisations

Organisations were identified through 5 international members lists: International Network of Agencies for Health Technology Assessment (INAHTA), Health Technology Assessment International, Red de Evaluación de Tecnologías Sanitarias de las Américas, Health Technology Assessment Asia and European Network for Health Technology Assessment (EUnetHTA) (as of April 2020), by cross– reference to organisations included in two previous HTA surveys of medical devices [10] and surrogate outcomes[19], and through expert recommendations provided by the project’s advisory group.Organisations were eligible if active, or closed within the last 3 years, organised HTA at a national level, defined by the World Health Organisation (WHO) as ‘institutionalized HTA’ [20], and included diagnostic tests within their remit of health technologies. To determine eligibility, the role and activities of each organisation were assessed using three sources of information: 1) information about each organisation on HTA network websites (e.g. INAHTA); 2) HTA organisation’s website, generally in an ‘About Us’ section or similar; and if both these approaches were unsuccessful 3) by web searches using the organisation’s name.

We excluded organisations whose remit did not include medical tests (e.g. All Wales Therapeutic and Toxicology Centre), but included agencies whose remit was less clearly stated. Organisations focussing entirely on evaluating population screening programmes were also excluded (e.g. UK National Screening Committee). We also excluded private research institutes, commercial organisations (e.g. manufacturers of health technologies), patient organisations, and university departments or other research groups undertaking HTA methods research, or producing HTAs not commissioned by an HTA organisation.

### Identifying methods and process guides

Initially, documents were eligible for examination if they were easily located from an organisation’s website and had the explicit aim of providing methodological guidance to reviewers undertaking HTAs for the organisation. We excluded methodology papers or journal articles unless they were clearly identified as representing the HTA organisation’s current methodology guidance. No language restrictions were applied. We restricted our examination to an organisation’s most current version of methods guidance.

Websites of included organisations were searched between March and July 2020 by visual inspection to locate relevant menus or page sections (e.g. ‘our methods’; ‘processes’). An automatic text translator was used for non–English language websites. If this process was unsuccessful, we used embedded search boxes with text strings: ‘guide’, ‘methods’, ‘methodology’, ‘process’, and ‘HTA’. We allocated a maximum of one hour searching time per HTA organisation to find relevant documents.

### Selecting organisations with detailed test–specific guidance

Our pragmatic approach aimed to focus on a detailed examination of organisations that were likely to have the most thorough methods guidance for carrying out HTAs of tests. We identified all organisations that had at least one methods chapter specific to test evaluation. From this sample we identified eligible organisations as those with the most detailed and informative methods guides and obtained any additional process documentation available on their websites.

### Extraction and Analysis

We collected data in two stages. In Stage 1 we performed an in–depth examination of selected organisations’ process and methods documents. All description relating to diagnostic tests was extracted verbatim into a piloted extraction form in the following domains: topic referral, prioritisation and scoping processes (pre–evidence review); identification, selection, appraisal and synthesis of evidence for clinical effectiveness reviews; methods for health economics evaluations; developing evidence into guidance; and stakeholder processes. A copy of the extraction form is provided in Supplementary file 1.

Each organisation was extracted independently by at least 2 researchers. Succinct summaries of each organisation’s approach were sent to a representative of each agency to validate and ensure the accuracy of our understanding. Responses were collated and discussed for inclusion. Six of seven organisations contacted agreed to have their responses published as part of our data extraction (Supplementary file 2). Approach to HTA of tests were summarised in the form of concordant and discordant themes across organisations and by in-depth discussion at team meetings.

In Stage 2 we performed a rapid interrogation of the methods guides from organisations without detailed HTA test methods guidance using the key themes generated in Stage 1. Methods documents were interrogated using common test–related terms, including “diagn”, “test”, “screen”, “detect”. For documents containing at least one test–related term, documents were assessed for presence and quantity of reporting for each theme. Duplicate independent extraction of Stage 2 organisations was performed when test–related terms were present in the absence of test–specific methods, and to resolve extraction queries. All data are presented descriptively.

## Results

Of 216 unique organisations, we included 41 with a remit for test evaluation and identified publicly available methods guidance (Figure 1). Experts recommended two non–HTA organisations (Grading of Recommendations, Assessment, Development and Evaluations (GRADE) [21] and WHO [22]) on the basis that they might provide important methodological innovations. The included organisations are listed in Supplementary Table 1.

**Figure 1:**
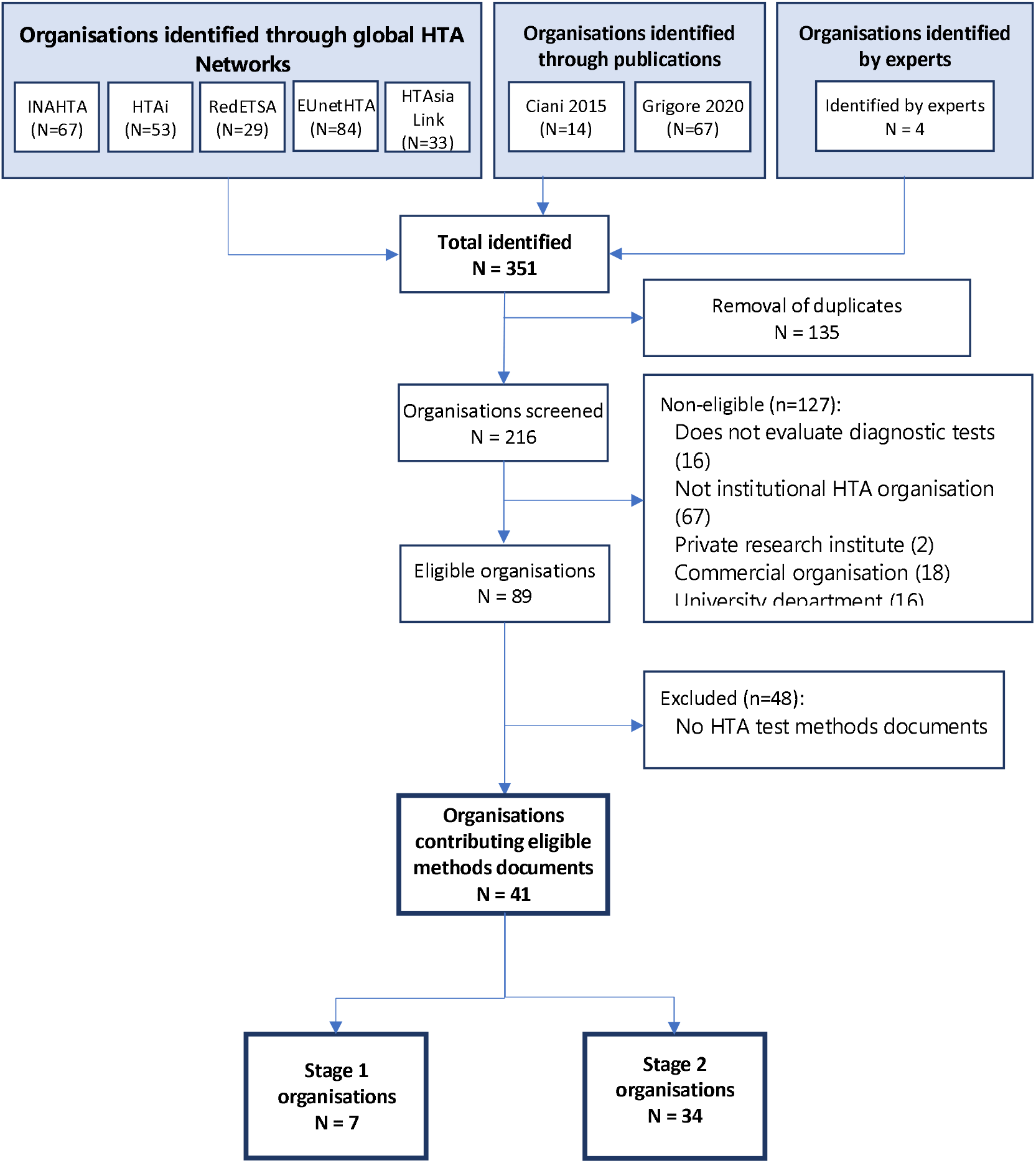
Identification of international HTA organisations and documentation.

### Stage 1: Key organisations with dedicated sections on test–specific methods

We identified 10 organisations with test specific guidance sections. Three of these were not considered in Stage 1 but rather in our analysis of organisations at Stage 2. The reason for this for two organisations was that test–specific sections were limited to an overview of test accuracy challenges (Gesundheit Österreich GmbH (GOG) and Agency for Care Effectiveness (ACE)). The EUnetHTA Core Model [23] was also deferred to Stage 2 because it is an international collaboration for development and dissemination of standardised methods for HTA [24] rather than an organisation undertaking HTA for a particular country and so strictly speaking not eligible.

The seven key organisations with test specific guidance sections were: AHRQ [15], Canadian Agency for Drugs and Technology in Health (CADTH) [25], Institut für Qualität und Wirtschaftlichkeit im Gesundheitswesen (IQWIG) [26], Medical Services Advisory Committee (MSAC) [27], NICE DAP [14], Statens beredning för medicinsk och social utärdering (SBU) [28], and Zorginstituut Nederland (ZIN) [29]. Supplementary Table 2 summarises the remit, capacity and sources we used for these seven organisations. In addition, representatives from six of seven included organisations provided feedback on our methods summaries (Supplementary file 2). Methods guides were last updated between December 2011 (NICE DAP) and May 2021 (MSAC), including three during the project (MSAC, IQWiG, SBU). Three organisations produced test–specific methods guides (NICE DAP, AHRQ, ZIN), one a guide for non–pharmacologic interventions (MSAC), and three test–specific guidance as chapters and/or dedicated appendices within generic HTA methods guidance documents (SBU, IQWiG, CADTH). Although CADTH do perform clinical effectiveness reviews, their publicly available methods guide is for health economic evaluation only.

### Thematic Analysis Findings

A summary of findings for Stage 1 organisations is provided in Table 2 and elaborated below.

**Table 1:** Summary of information on main themes in each key organisation

|  | AHRQ | CADTH | IQWiG | MSAC | NICE DAP | SBU | ZIN |
| --- | --- | --- | --- | --- | --- | --- | --- |
| Defining Review Question | PICOTS<br>Population;<br>Intervention (Index test); Comparator;<br>Outcome; Timing;<br>Setting (and study design) | Makes test specific considerations such as the importance of the diagnostic setting and place of the test in the management pathway | Follows international standards of evidence-based medicine without overt tailoring to diagnostic tests | All aspects of PICO contain test-specific considerations | All aspects of PICO contain test-specific considerations | PICO with modification to PIRO for test accuracy studies | PICO (formulated on basis of claims of test): Patient and setting, I=test-plus-treatment strategy, C=comparative test-plus-treatment strategy, O=outcomes relating to patient health.<br>Focus on defining role of test |
| Defining Claims of Tests | Use of a framework to systematically identify claims – Analytic framework:<br>— designed to generate questions that direct selection of evidence<br>— to be used during scoping<br>— uses causal pathway approach, with graphic | Not reported | No test specific guidance provided | Use of a framework to systematically identify claims – Analytic framework:<br>— designed to generate questions that direct selection of evidence<br>— to be used during scoping<br>— uses causal pathway approach, with graphic | No structure reported. A key part of scope preparation is to identify any outcome that could be impacted on by introducing the test in the NHS | Not reported | Use of a comparative analysis framework to systematically identify claims:<br>— designed to generate questions that direct selection of evidence<br>— uses test-plus-treatment mapping |
| Outcome prioritisation | Yes | Not reported | Not reported | Yes | No, comprehensive approach | Not reported | Yes |
| Claims inform ER or HEE methods | ER | Not reported | Not reported | ER and HEE | Unclear | Not reported | ER |
| Evidence preference | Direct evidence preferred but valid and applicable evidence unlikely to be found. Indirect evidence using narrative evidence linkage, with some methods | Direct evidence preferred. Not explicitly addressed as methods guide is for HEE review only | Direct evidence preferred with a focus on RCT. Indirect may be considered, but methods not provided. Research can be commissioned if direct evidence not available | Direct evidence preferred but valid and applicable evidence unlikely to be found. Indirect evidence using narrative evidence linkage, with extensive methods, including GRADE adaption | Direct evidence preferred but valid and applicable evidence unlikely to be found. Indirect evidence using evidence linkage formal modelling, no methods reported | Direct evidence preferred. Indirect may be considered, but methods not provided. The relative strength and importance of evidence is assessed using the GRADE framework for tests, using an approach modified by SBU | Direct evidence preferred but valid and applicable evidence unlikely to be found and may not be necessary. Indirect evidence using evidence linkage, no methods reported. EBRO classification is recommended to determine the level of evidence. GRADE is referred to in appendix of ZIN's methods guide |
| Searching for evidence | Accepted methods for systematic reviews, with specific tailoring for tests. Emphasis for tests includes: caution not to rely on search filters alone or on controlled vocabulary (subject headings) alone; advice to search in multiple databases/ locations with tailored strategies with advice on designing search terms | No publicly available methods guidelines for performing a clinical effectiveness review for CADTH HTA products. Mention is made of: including a multidisciplinary team and; peer-review by external clinical, economic, and methodological | For searching, international standards of EBM are applied. No special considerations for tests mentioned, except noting uncertainty about search filters for accuracy studies | Accepted methods for systematic reviews, with additional test methods including: multiple iterative searches, regardless of whether linked evidence approach is required; separate searches for treatment-related outcomes and for safety outcomes. Advice on searches: caution on search filters; additional | Search guidance for test accuracy evidence and other outcomes. Recommend different strategies for less complexities of diagnostic technology. Recommendations on: avoiding diagnostic search filters; using specialist database searches; reference chasing; and | Clinical evidence is identified using general methods for systematic reviews, with no specific advice for diagnostic questions other than to use the 'PIRO' to create the search strategy | Systematic search methods mentioned, but no technical guidance specific to test evaluations |
|  |  | experts and internal CADTH staff |  | searching in clinical trial registers, manual check reference lists; guidance for searching for 'value of knowing' claims | sometimes abstract searches. If no end to end studies, additional searches for 'linked evidence' |  |  |
| Quality assessment | Use of validated criteria recommended. For DTA studies recommend QUADAS-2. For studies using tests as interventions, quality should not differ greatly to those used for intervention studies. Quality of other study designs not mentioned. Applicability includes consideration of spectrum effects, roles of tests, shifting routine care, evolving versions of tests | No reference to specific methods for clinical effectiveness reviews of tests | International standards of EBM are used and referenced: QUADAS-2 for diagnostic accuracy studies; PROBAST for prognostic studies; intervention studies standard recommended EBM methods | Table of validated tools provided for 11 different study designs including methods suitable for therapeutic technologies. Test-specific tools accuracy tools recommendations are: QUADAS-2; STARD; ACCE for genetic studies and QUIPS for prognostic studies. Detailed discussion on the ways in which the generalisability of various study designs may falter | Quality assessment approaches are detailed, particularly for test accuracy (use of STARD and QUADAS), but also for some other included study types. A list of types of bias applicable to test accuracy evidence is presented | Recommend QUADAS-2 instrument with appropriate tailoring for diagnostic accuracy. No guidance on how to incorporate quality assessments into review | Methodological quality assessment using QUADAS-2 is mentioned but no technical guidance specific to test evaluations or, more generally, to health interventions, is provided in the reviewed documents |
| MA guidance | Detailed guidance on MA where there is a reference standard, including Cochrane DTA Handbook. Provides options for summarizing medical test performance in the absence of a "gold standard" | Directed towards methods in existing systematic review and meta-analysis articles/Cochrane DTA handbook | Synthesis using international standards of EBM, with specific meta-analysis methods for DTA | Detailed methods on DTA meta-analysis including reference to internationally recognised standard methods. Advice on direct test-health outcomes and safety/harms outcomes | References to Cochrane DTA Handbook. Extensive advice on synthesis of test accuracy studies, with guidance for other study designs. Guidance provided about meta-analysis | Only test specific methods are for DTA meta-analysis. Heterogeneity is 'the rule rather than the exception' Direction to discuss consequences of FNs and FPs | Refers to methods in Cochrane DTA Handbook for assessment of test accuracy, including meta-analysis |
|  | Principles of GRADE for interventions can be used or adapted |  |  |  | including when meta-analysis would not be considered appropriate |  |  |
| HEE done | No | Yes | Optional | Yes | Yes | Yes | Optional |
| HEE methods | N/A | Health economic modelling | Health economic modelling | SR of economic studies and health economic modelling | SR of economic studies and health economic modelling | SR of economic studies and health economic modelling | Health economic modelling |
| Tailoring of HEE methods to tests | N/A | Good general guidance with pointers to specific issues when evaluating tests especially in specific guidance on companion diagnostics | Good general guidance on how to conduct an economic evaluation. No tailoring | Most detailed tailoring encountered but still relies heavily on general guidance on how to conduct cost-effectiveness and cost-minimisation analyses | Pointers to specific issues needing to be considered when evaluating tests. Limited detail however | Good general guidance on how to conduct an economic evaluation. No tailoring | Good general guidance on how to conduct an economic evaluation. Little tailoring. Refers to NICE DAP for more detail |
| Role of modelling | Modelling used for enhancing the interpretation of systematic reviews of diagnostic test accuracy | Modelling used as part of economic evaluation. Tool by which evidence linkage between test accuracy and patient outcome is achieved | Modelling used as part of economic evaluation. No reference to evidence linkage or use of modelling to achieve this | Modelling used as part of economic evaluation. Main approach to evidence linkage is to create narrative chain. Emphasised that modelling structure should reflect this narrative chain | Modelling used as part of economic evaluation. Tool by which evidence linkage between test accuracy and patient outcome is achieved | Modelling used as part of economic evaluation. No reference to evidence linkage or use of modelling to achieve this | Modelling used as part of economic evaluation. "Comparative analysis framework for indirect evidence" is the main approach to evidence linkage with no mention of modelling |

Abbreviations and notes:

|  | AHRQ | CADTH | IQWiG | MSAC | NICE DAP | SBU | ZIN |
| --- | --- | --- | --- | --- | --- | --- | --- |
| Canadian Agency for Drugs and Technology in Health; DTA – diagnostic test accuracy; EBM – evidence-based medicine; EBRO – Evidence-based Guideline Development in the Netherlands; ER – Evidence review; FN – false negative; FP – false positive; GRADE – Grading of Recommendations, Assessment, Development and Evaluations; HEE – health economic evaluation; HTA – health technology assessment; IQWiG – Institut für Qualität und Wirtschaftlichkeit im Gesundheitswesen; MA – meta analysis; MSAC – Medical Services Advisory Committee; N/A – not applicable; NICE DAP – National Institute for Health and Care Excellence Diagnostics Assessment Programme; PICO – population, intervention, comparator, outcome; PIRO – population, intervention, reference standard, outcome; PROSPERO - International Prospective Register of Systematic Reviews; PROBAST – Prediction model Risk Of Bias ASessment Tool; QUADAS – Quality Assessment of Diagnostic Accuracy Studies; RCT – randomised controlled trial; SBU - Statens beredning för medicinsk och social utvärdering; STARD – STANDards for reporting of Diagnostic accuracy; ZIN Zorginstituut Nederland |  |  |  |  |  |  |  |

**Table 2:**
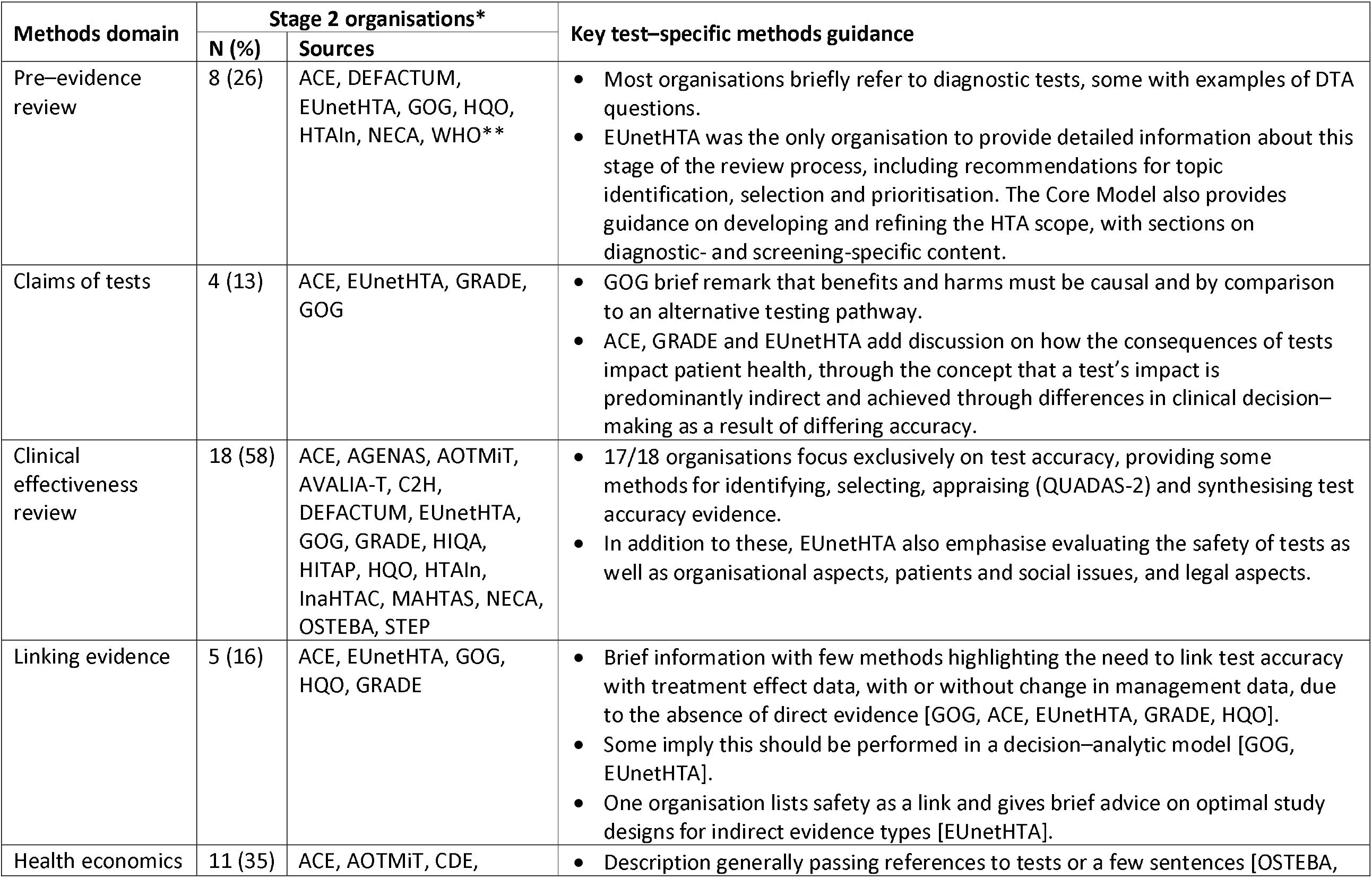
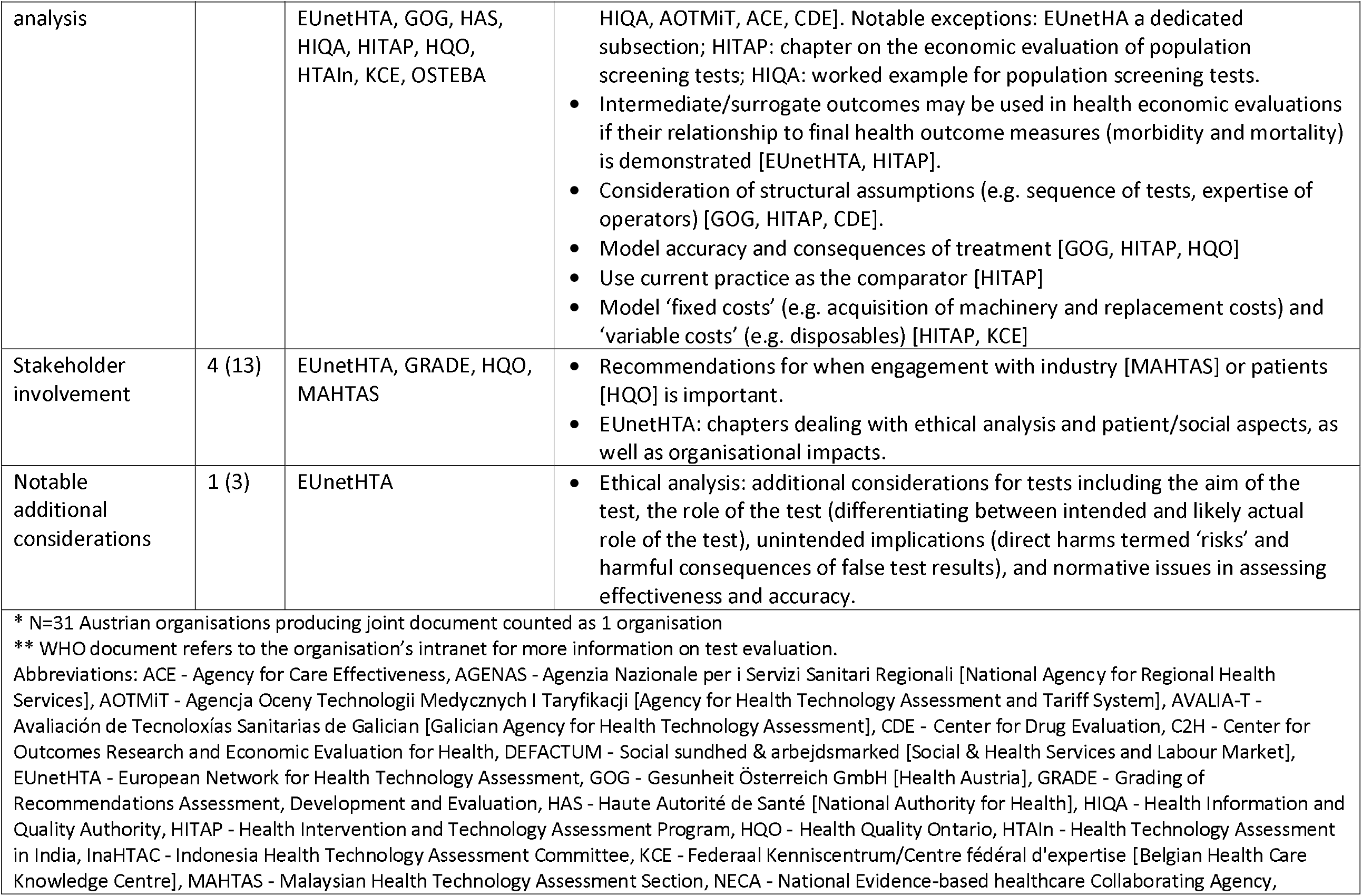
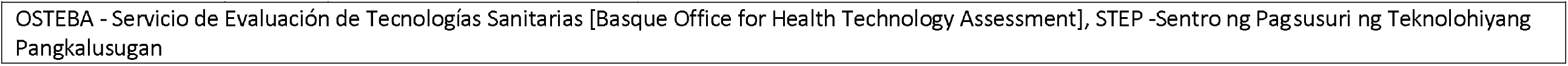
Summary of information on organisations examined at Stage 2

#### Claims of tests

Establishing how a study test might be clinically effective is discussed to varying degrees by all organisations, who share the same philosophy of the primacy of patient health outcomes, and that tests impact these outcomes mainly indirectly through the consequences of test results. A shared key principle arising from this philosophy is that evidence of accuracy is on its own insufficient to demonstrate the test’s clinical effectiveness, since test results must also demonstrate a change to patient management, which is shown to change health outcomes.

The organisations differ, however, in how the claims of tests are used to guide the HTA process (Figure 2). Both AHRQ and MSAC use a structure to construct the claims of a test in a systematic manner, which is achieved using a graphic to map out the main components of patient care from testing to health outcomes, for each comparative management pathway. Both are adaptations of the USPSTF’s analytic framework [30], where a causal pathway approach is used to identify key differences in the care a patient will receive as a result of using different tests, which in turn generates review questions that direct the selection of evidence. ZIN use a ‘comparative analysis framework’, which employs a similar approach of mapping out ‘test–plus–treatment’ pathways, and discerning important differences and consequences to patient health as a result of changing the test (ZIN).

**Figure 2:**
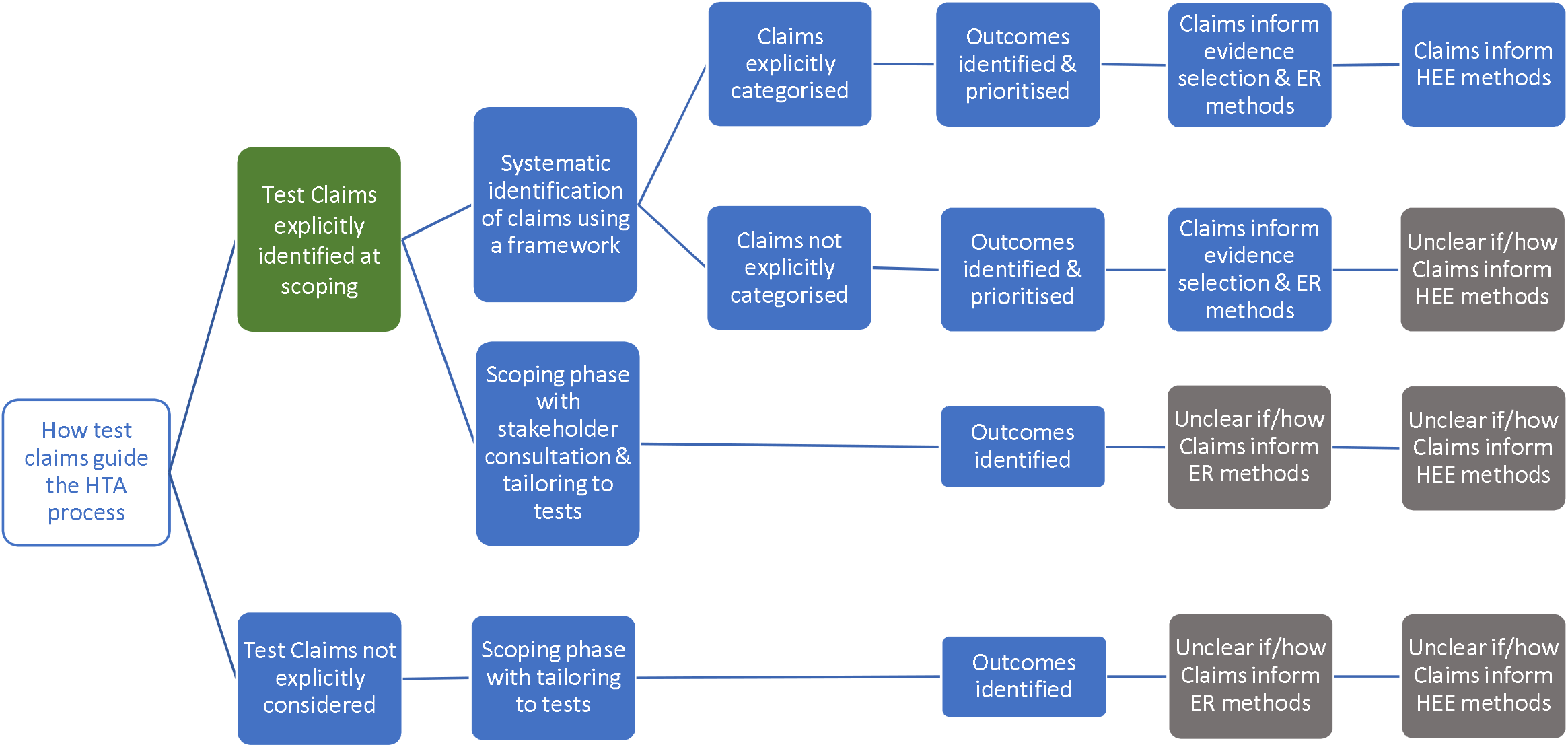
Stage 1 organisation approaches to defining the way in which a diagnostic test claims to impact on patient health. Green cells identify the majority view, grey cells indicate uncertainty in approach, and blue cells illustrate specifically reported approaches to identifying and using test claims in the HTA process. ER – Effectiveness Review; HEE – Health Economics Evaluation; HTA – Health Technology Assessment.

AHRQ, MSAC and ZIN use the claims framework to identify and prioritise which outcomes should be evaluated as part of the HTA, and as such this process forms a central part of the scoping phase for each organisation. MSAC provides detailed example frameworks with associated review questions, organised according to a categorisation of claims, as well as conditions when it is appropriate to truncate the framework for a claim of non-inferiority (no difference in health outcomes). Further detailed guidance is also given for different diagnostic comparisons (i.e. replacement, add–on, triage), as well as for various types of test, for example multifactorial algorithms and machine learning/artificial intelligence tests.

NICE DAP conduct a thorough scoping process, in which outcomes are identified using pathways analysis and extensive stakeholder consultation, however they do not actively prioritise those outcomes. Outcomes prioritisation was not discussed by the remaining organisations (SBU, CADTH, IQWiG), however all seven organisations use test–specific adaptations of the Population Intervention Comparator Outcome (PICO) structure, emphasising test comparison, and all cite some additional requirements for tests, most commonly the role of the study test within existing care pathway. All organisations involve multiple stakeholders groups during question formulation, including clinical experts and patient/user representatives.

The degree to which the type of claim informs the method by which it is evaluated in an HTA was also found to vary, with an integrated approach described by ZIN, AHRQ and most clearly and thoroughly by MSAC. In all three, claims underlie the search for, selection and interpretation of evidence; AHRQ for example uses claims to inform quality assessment, particularly applicability, while organisations in whom claims do not appear to inform methods tend to use the PICO for this purpose (NICE DAP, SBU). For MSAC, a test’s claims underpin all subsequent HTA methods, from searching through to analysis, synthesis and health economics. To achieve this, MSAC have developed a ‘hierarchy of claims’ (MSAC pp26-27), in which a health claim must always be made (that the ‘new’ test results in superior or non–inferior patient health) but which also allows for claims of non–inferiority to be accompanied by additional ‘non–health’ claims, which MSAC terms ‘value of knowing’ (MSAC p18). The principal claim should relate to health, and although this is explicitly derived from the expected comparative accuracy of tests, methods to demonstrate non-inferiority in test accuracy are not provided. We did note that MSAC’s advice for analysing comparative accuracy was removed from their 2017 guidance [31].

#### Direct and Indirect evidence for the clinical effectiveness of tests

Figure 3 summarises the approaches Stage 1 organisations advocated for using direct and/or indirect evidence to investigate the effectiveness of diagnostic tests. All seven organisations state a clear preference for ‘direct’ evidence, also referred to as ‘end–to–end’ evidence (NICE DAP), ‘head–to– head effectiveness’ (AHRQ) or ‘direct test to health outcomes evidence’ (MSAC). This was consistently defined as comparisons of patient health outcomes following differing tests or testing strategies within a single primary study. The most common example given is of RCTs that randomise patients between tests and follow them through subsequent clinical management, after which health outcomes are measured.

**Figure 3:**
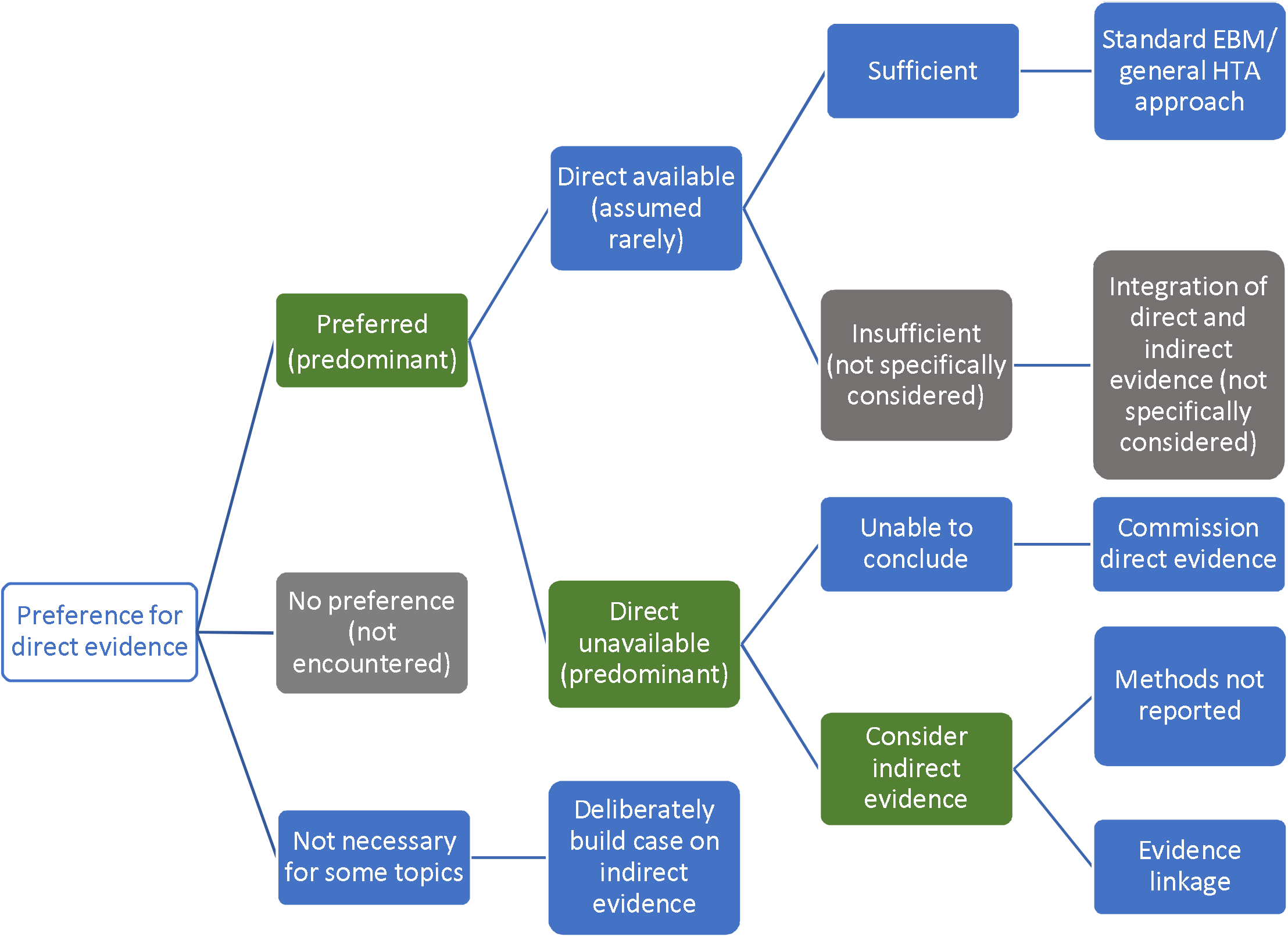
Stage 1 organisation approaches to using direct and indirect evidence to evaluate the effectiveness of diagnostic tests. Green cells identify the majority view, grey cells are not specifically mentioned but are plausible approaches, and blue cells illustrate specifically reported approaches to using direct and indirect evidence.

We found little test–specific guidance for how to assess and synthesise direct evidence. The most common recommendation is to follow international EBM methods for appraising and synthesising RCT evidence of treatment interventions (AHRQ, MSAC, ZIN, IQWiG, NICE DAP), including use of GRADE to assess the certainty/strength of the body of evidence (MSAC, SBU, AHRQ, ZIN). MSAC do supplement this treatment intervention method with three key additional components: examining the applicability of direct evidence, for which detailed guidance is given; a requirement to report how direct test–health outcomes evidence has been constructed; and the need to present evidence of test–related harms (both direct procedural harms and harms following subsequent management decisions) as a separate section of synthesis (MSAC, pp99-115).

There was a majority view that relying on direct evidence to inform an HTA is likely to be problematic, since these ‘test–plus–treatment RCTs’^33^ are uncommon (NICE DAP, MSAC, AHRQ, SBU, ZIN) and have particular challenges to their reliability and generalisability (AHRQ, MSAC, ZIN).

#### Handling indirect evidence

There does not appear to be a clear consensus on how to proceed when direct evidence is unavailable, insufficient or unnecessary (Figure 3). Some organisations suggest that their main approach is to use direct evidence (IQWiG, SBU), and commission primary research when it is not available (IQWiG). Both SBU and IQWiG suggest they may also use ‘indirect’ evidence, such as evidence of a test’s accuracy (or any other outcome that is intermediate between a test and health outcome), however neither organisation’s guidance provides explicit methods for how this should be used to determine clinical effectiveness.

The main alternative view to the use of direct evidence is “evidence linkage” (NICE DAP, AHRQ, MSAC, ZIN): test accuracy evidence (the proportion of individuals correctly classified as true positive and true negative) is linked with effectiveness evidence (the effects of appropriate treatment) and compared across different test-treatment strategies to provide an estimate of the likely overall effect of the test on a health outcome.

Two types of evidence linkage are identified, narrative and formal modelling. MSAC provides a detailed explanation of how to create a narrative chain of argument showing how different bodies of evidence confirm or challenge the likelihood that improvements in accuracy will lead to changes in clinical management and subsequently clinical outcomes. For MSAC, AHRQ and ZIN, methods to select and assess indirect evidence are explicitly linked to a test’s claims, which enables an explicit examination of the adequacy of ‘links’ between different types of evidence, such as between a test accuracy study and an impact to clinical decision–making study. This ‘transitivity’ (MSAC) or ‘transferability’ (AHRQ) of evidence forms a key method in both organisations, to which AHRQ add the recommendation to apply GRADE to each of these links in addition to grading the overall body of evidence, although they identify a lack of methods for doing so. MSAC outline specific adjustments to the standard GRADE approach for each of their three pre–specified evidence links. ZIN provide an accessible stepped process for developing and conducting a comparative analysis (ZIN pp28–29), while MSAC also present useful suggestions for producing an informative narrative synthesis, including explaining the summative benefits and harms for a hypothetical population from the point of undergoing testing.

The alternative approach to narrative evidence linkage is formal modelling of links between accuracy and effectiveness evidence (NICE DAP). There is limited detail on how this might be achieved and it is implied that methods for economic modelling, such as decision-analytic modelling, can be simply extended to create a linked evidence model of effectiveness and cost-effectiveness.

Aside from the narrative summary guidance provided by MSAC, methods for synthesising indirect evidence were exclusively restricted to test accuracy meta–analysis (NICE DAP, SBU, IQWiG, CADTH, AHRQ). We also did not find reference to specific methods for judging the adequacy of linked cases, beyond notions of applicability, including how to deal with inconsistencies in the evidence base between different study types. Similarly, we did not find guidance for how to resolve inconsistencies in findings between direct and indirect evidence, nor how to assess the strength of such a mixed evidence base.

#### Searching for evidence

All seven organisations acknowledge the importance of conducting comprehensive and systematic searches and presenting the results in a clear and transparent way. MSAC, NICE DAP and AHRQ state that main databases and trial registries (e.g. MEDLINE, EMBASE), should be supplemented with other databases, reference checking and contacting experts to identify additional titles and unpublished data. Some differences were noted. There is diversity concerning use of diagnostic search filters with NICE DAP, IQWiG, AHRQ and MSAC advising caution against use, whilst AHRQ recommend further investigation of the option. Concerning search strategy NICE DAP recommend a 2-step approach first searching for direct evidence and then, if such evidence is not available or sufficient, conducting additional searches to inform a linked evidence analysis. MSAC set out a more iterative approach including the need for multiple searches depending on the test’s health claim and availability of direct ‘test–health outcomes’ evidence. Notable additions include systematic and targeted searches to supplement direct evidence when it is found to be insufficient to answer the review question, to: assess the safety of tests; address each evidential link identified by the assessment framework, including separate searches for each change in management occurring as a result of the test; or to find evidence for exploring any ‘value of knowing’ claims.

#### Quality assessment

The importance of assessing the risk of bias (internal validity) and applicability (external validity) of the identified evidence in HTAs of tests is acknowledged by all key organisations with most focus being on the approach for test accuracy. The level of guidance provided varies. Risk of bias should be assessed using validated tools, most commonly (AHRQ, IQWiG, MSAC, NICE DAP, SBU, ZIN) Quality Assessment on Diagnostic Accuracy Studies-2 (QUADAS-2) is recommended for assessing diagnostic accuracy studies. NICE DAP and MSAC also suggest the use of STARD, without discussing the advantages and disadvantages of this approach. MSAC provides a valuable inventory of quality assessment tools for a range of different study designs, including RCTs and observational studies (MSAC pp 251-53), some of which are also recommended by other key organisations. For designs other than accuracy, the general assumption is that no adaptation of tools is required when the focus is tests, as in the case of RCT: “For trials of tests with clinical outcomes, criteria should not differ greatly from those used for rating the quality of intervention studies” (AHRQ). All key organisations recommend assessing the applicability of evidence, specific to the country and setting in which the test is to be used, and to review questions. Again, the level of guidance varies, from general recommendations related to the methodological quality assessment of individual studies (IQWiG) to more detailed treatment of the topic for all stages of the HTA process (MSAC, NICE DAP and AHRQ).

#### Health economic evaluation

Six of the key organisations assess cost-effectiveness of tests although with different emphasis. MSAC is explicit that it is one part of the considerations, clinical need, comparative health gain and predicted use in practice and financial impact being the others. IQWIG and ZIN imply that, although helpful, economic analysis is not always necessary, while at the other end of this spectrum CADTH and NICE DAP place emphasis on it. Where cost-effectiveness is used, the main approaches are to systematically review existing economic evaluations and to use model-based economic evaluation often estimating cost per QALY (quality-adjusted life year). They all provide good guidance on the general approach to conducting model-based evaluations, which could be applied irrespective of the technology under consideration. However, there is very little guidance on how general approaches need to be adapted to particular demands of evaluating tests, beyond offering pointers in the case of NICE DAP and CADTH. This gives the impression that there are a limited number of challenges which can be easily overcome. MSAC offers the greatest amount of detail but still relies heavily on general guidance on how to conduct cost-effectiveness and cost-minimisation analyses.

#### Additional contributions

We identified distinct additional contributions from all seven organisations, summarised in Supplementary Table 3. Notable methods include a framework to systematically identify the ethical aspects of any health technology [32] (although no specific tailoring for tests) (SBU), a framework for assessing technical modifications of tests from ZIN, and guidance for specific types of diagnostic test, including genetic tests from AHRQ and MSAC, codependent tests from CADTH and MSAC) and artificial intelligence/machine learning tests from MSAC.

### Stage 2: Systematic check of test–specific methods in other organisations

We performed a rapid interrogation of manuals produced by 34 organisations not considered in Stage 1 (Figure 1). One document was jointly produced by four Austrian organisations (GOG, Austrian Institute for Health Technology Assessment, University for Health Sciences, Medical

Informatics and Technology, Donau-Universität Krems) so there were 31 unique sources the findings from which are summarised in Table 2. Two organisations for whom we did not find methods manuals, referred to the EUnetHTA Core Model methods, considered in Stage 2, on their websites: Finnish Coordinating Center for Health Technology Assessment [https://www.ppshp.fi/Tutkimus-ja-opetus/FinCCHTA/Sivut/HTA.aspx] and Ministry of Health of the Czech Republic [https://www.mzcr.cz/category/metodiky-a-stanoviska/].

Most Stage 2 organisations referred to tests in at least one document, however these tended to be short with few definitive methods. When present, methods were commonly limited to evaluating and synthesising test accuracy studies (Avaliación de Tecnoloxías Sanitarias de Galician, Social sundhed & arbejdsmarked, Indonesia Health Technology Assessment Committee, Health Information and Quality Authority, Agenzia Nazionale per i Servizi Sanitari Regionali, GOG, Health Technology Assessment in India, Malaysian Health Technology Assessment Section, Sentro ng Pagsusuri ng Teknolohiyang Pangkalusugan, WHO). There were some exceptions. Detailed guidance was provided by EUnetHTA ‘Core Model (version 3)’ [23] and associated documents provide test– specific methods for all components of an HTA, from scoping through to clinical and cost– effectiveness.

Stage 2 organisations rarely discussed how to discern a test’s claims. Those that did shared the same philosophy of a test’s predominantly indirect impact, achieved most commonly through differences in clinical decision–making as a result of differing accuracy (ACE, GRADE, EUnetHTA). Test–specific methods were most commonly located in clinical effectiveness review sections, which were always focussed on test accuracy, with no tailoring of methods to evaluate direct evidence. A few organisations provided descriptions of linking evidence (ACE, EUnetHTA, GOG, Health Quality Ontario, GRADE), which may include decision–analytic modelling (GOG, EUnetHTA). Like MSAC, EUnetHTA emphasise the importance of evaluating safety as a type of outcome in its own right, distinguishing ‘reduced risk’ (direct test harms) and ‘increased safety’ (as a result of better accuracy). EUnetHTA are the only organisation to distinguish safety from clinical effectiveness evidence, and although EUnetHTA highlight a likely overlap between the two, no methods are suggested for how to overcome the possibility of double–counting adverse consequences of inaccuracy. Alongside accuracy and diagnostic/therapeutic impact, EUnetHTA also define ‘other patient outcome[s]’, such as knowledge and increased autonomy, which appear to be analogous to MSAC’s ‘value of knowing’ outcomes. While EUnetHTA do not prioritise outcomes per se, they do describe the process of identifying the most important consequences (and hence outcomes) as ‘value–decisions’ that demand transparency in how they are made.

Methods for health economic analysis infrequently referred to tests as the subject of evaluation and did not provide additional methods to those found in our Stage 1 assessment. EUnetHTA provide test-specific guidance for ethics analysis and for assessing the broader issues of organisational aspects, patients and social issues, and legal aspects, methods cited by MSAC as a key source for undertaking these investigations. Ethical analysis in particular constitutes a notable development in comparison to key organisations in Stage 1, detailing additional test–specific considerations including the aim of the test, the role of the test (differentiating between intended and likely actual role of the test), unintended implications (direct harms termed ‘risks’ and harmful consequences of false test results), and normative issues in assessing effectiveness and accuracy [23].

## Discussion

We present a first review of HTA organisation methods for performing HTAs of tests.

### Main findings

In general, few test–specific methods are provided by HTA organisations that evaluate diagnostic tests, and when present these methods predominantly concern the evaluation of test accuracy evidence. A small number of organisations do provide more detailed tailoring of HTA methods for tests.

Within these key organisations, there is consistency in many respects. One is the need for the complexity of the diagnostic process to be fully incorporated into question definition, which favours the pre–eminence of patient health outcomes. Approaches to the searching, quality assessment and meta-analysis of test accuracy studies is another. With the exception of dealing with test accuracy evidence, all HTA organisations tend to defer to general HTA approaches, designed to assess pharmaceuticals, rather than approaches specific to the HTA of tests.

There is also divergence. We found critical differences in methods used to identify the mechanisms by which a diagnostic intervention may impact health outcomes. Although prior methods research demonstrates these claims of tests are composed of numerous, interdependent mechanisms that are intrinsically based on the clinical setting [2], most HTA organisations we examined did not provide methods for identifying the claims of the test intervention. While some key organisations provide an explicit method, consisting of a structured a priori framework, others rely on the scoping process to discover – but not prioritise – a test’s claims, and hence the evidence and outcomes required to assess clinical effectiveness. Here, the current best practice is showcased most clearly by MSAC who embed test claims as a foundation, informing the ensuing methods for evaluating clinical evidence and cost–effectiveness. Regardless of approach, all organisations frame differences in accuracy as the principal claim of a test’s impact to patient health.

Although all HTA groups recognise the primacy of direct evidence of the effect of a test on health outcomes, there are a range of views on how to proceed in its absence. At one extreme the emphasis is on encouraging the generation of direct evidence; at the other is an openness to all forms of evidence. Evidence linkage is a prominent solution, varying from narrative linkage to formal modelling, though it is unclear whether there is consistency in these approaches because most organisations do not clearly define them, and provide limited detail for how they should be performed. MSAC is the notable exception, describing a process by which evidence is sought with respect to each identified test claim, and a narrative chain of argument is created showing how improvements in test performance will be translated into improved health outcome, also incorporating harm and value of knowing.

### Strengths and weaknesses

Although this methods review is subjective, we have attempted to maximise objectivity by working to an a priori protocol, performing all steps in duplicate, and offering key (Stage 1) organisations the opportunity to check the face validity of summaries. We were exhaustive in our efforts to identify as many HTA organisations as possible, and to give equal opportunity for all of these to be considered eligible for detailed scrutiny as key organisations. We recognise a limitation of our research is an assumption that guidance reflects actual practice in relation to HTA of tests, whereas the reality may be that this guidance is not followed or is impractical to adopt by the organisations producing the individual HTAs. Further, our comparison of international HTA approaches may not fully account for the different contexts in which HTA is undertaken across different organisations. For example, some organisations have a remit to make recommendations for policy and reimbursement whereas others have a remit for evidence synthesis only. These contextual factors could have an impact on the level of detail and type of guidance offered.

### Findings in context

We are not aware of any other reviews that have examined methods for undertaking all phases of an HTA, for determining the clinical and cost–effectiveness of diagnostic tests. Our findings do mirror those of a previous comprehensive assessment of test focused HTAs, which reviewed methods used to synthesise evidence informing economic decision models for HTAs of tests [17]. A key finding concerned the lack of a clear framework to identify and incorporate evidence into health economic models of tests. Despite being conducted five years later, our review found very few health economics methods were tailored specifically to tests, suggesting that this ‘gap’ between clinical and cost–effectiveness may still remain. Further, our review suggests this gap may originate in failings to clearly delineate test claims at the outset of the HTA process.

### Recommendations

For practice, we found some good materials which would be useful for guiding an organisation new to HTAs of tests. As a comprehensive exposition of all aspects, we would particularly recommend the guidance by MSAC [27].

For research, aspects of divergence reflect areas where there is not yet consensus amongst the HTA community, and so indicate opportunities for methodological research. We would highlight further development of evidence linkage methods, particularly for managing and interpreting the heterogeneity of an indirect and linked evidence base. Another specific uncertainty concerns whether the evidence linkage achieved by developing a health economic model alone is a sufficient substitute for narrative evidence linkage, followed by economic model development as required. We also noted absence of advice on how to evaluate non–inferiority, with no guidance on how to identify, summarise and interpret comparative accuracy studies, which should be the ideal studies to assess non-inferiority [33]. A new tool, QUADAS-C, has recently been developed to specifically address risk of bias in comparative accuracy studies [34], and may be helpful in future guidance and updates. In addition, we would question whether sufficient attention has so far been paid to the complexities of identifying and interpreting direct evidence, and how a judgement is made that this evidence is sufficient or insufficient for decision–making. Methods reviews of test–treatment RCTs have shown these studies are more susceptible to particular methodological challenges than RCTs of medicines [35], suggesting that appraising their quality (both internal and external validity) may not be served well by the application of tools developed for treatment trials. Importantly, current guidance is silent on how to resolve inconsistencies between direct (end to end) evidence and indirect (linked) evidence (where both have been undertaken), which as MSAC note is the most likely scenario faced by reviewers of tests.

## Conclusion

This methodological review gives an indication of the global state of the art with respect to HTA of tests. Our study reveals many important outstanding challenges, not least the general need for more extensive and detailed guidance outlining methods that are tailored specifically to evaluate diagnostic tests. Although the evaluation and synthesis of accuracy evidence is the most comprehensively and commonly provided methods topic, there is a need to incorporate more recent innovations, such as approaches for incorporating and interpreting comparative evidence of test accuracy. Although all HTA organisations are clearly focused on evaluating the benefits and harms of healthcare technologies to patient health, we found there is often a hiatus in methods guidance between evaluating diagnostic accuracy and assessing the end clinical effectiveness of diagnostic interventions. Organisations providing the most detailed guidance in this area tended to be those that incorporated the clearest methods for identifying and understanding how tests impact on patient health. We believe this observation underscores the centrality of test claims as a method for explicitly informing the selection, identification, interpretation and synthesis of evidence used to deduce the clinical effectiveness of tests.

There has been a tendency in the past to de-prioritise HTA of tests, and methods development relative to HTA of pharmacological interventions. However, the prominence of tests among health innovations (genetic testing and artificial intelligence applied to imaging) and the rapidly escalating costs of new tests argues that now is the time for priority to be given to methods development in this area. The recent confusion and uncertainty around evaluation of tests and testing strategies for COVID–19 reveals just how frail the current systems are for sanctioning the uptake of new tests into healthcare. The origins of this lie substantially in the general uncertainty of how the HTA process for tests can best answer the question “What constitutes a good enough test”, which this paper explores.

## Data Availability

All data produced in the present study are available upon reasonable request to the authors

## Acknowledgements

Representatives of HTA organisations for their useful feedback: Christine Chang and Craig Umscheid, on behalf of the Agency for Healthcare Research and Quality; Andrew Mitchell, on behalf of the Medical Services Advisory Committee; Joanne Kim and Cody Black, on behalf of the Canadian Agency for Drugs and Technologies in Health; Stefan Sauerland on behalf of Institut für Qualität und Wirtschaftlichkeit im Gesundheitswesen; Thomas Walker on behalf of the National Institute for Health and Care Excellence; and Susanna Axelsson, Jenny Odeberg and Naama Kenan Modén, on behalf of Statens beredning för medicinsk och social utvärdering. Also Bogdan Grigore for his list of HTA organisations. Finally the MRC TEST Advisory Group for discussion of methods approach and comment on manuscript: Patrick Bossuyt, Katherine Payne, Bethany Shinkins, Thomas Walker, Pauline Beattie, Samuel Schumacher, Francis Moussy, Mercedez Gonzalez Perez.

## References

1. Fryback DG, Thornbury JR. The Efficacy of Diagnostic Imaging. Med Decis Making 1991;11:88–94.

2. Ferrante di Ruffano L, Hyde CJ, McCaffery KJ, Bossuyt PMM, Deeks JJ. Assessing the Value of Diagnostic Tests: A Framework for Designing and Evaluating Trials. BMJ 2012;344:e686.

3. Bossuyt PM, Lijmer JG. Traditional Health Outcomes in the Evaluation of Diagnostic Tests. Acad Radiol 1999;6 Suppl 1:S77-80; discussion S3-4.

4. Fineberg HV. Evaluation of Computed Tomography: Achievement and Challenge. AJR Am J Roentgenol 1978;131:1–4.

5. Schünemann HJ, Oxman AD, Brozek J, et al. Grading Quality of Evidence and Strength of Recommendations for Diagnostic Tests and Strategies. BMJ 2008;336:1106–10.

6. Ferrante di Ruffano L, Davenport C, Eisinga A, Hyde C, Deeks JJ. A Capture-Recapture Analysis Demonstrated That Randomized Controlled Trials Evaluating the Impact of Diagnostic Tests on Patient Outcomes Are Rare. Journal of Clinical Epidemiology 2012;65:282–7.

7. Merlin T, Lehman S, Hiller JE, Ryan P. The “Linked Evidence Approach” to Assess Medical Tests: A Critical Analysis. Int J Technol Assess Health Care 2013;29:343–50.

8. Sutton AJ, Cooper NJ, Goodacre S, Stevenson M. Integration of Meta-Analysis and Economic Decision Modeling for Evaluating Diagnostic Tests. Med Decis Making 2008;28:650–67.

9. Wilsdon TS, A. A Comparative Analysis of the Role and Impact of Health Technology Assessment. London: UK: Charles River Associates 2011

10. Ciani O, Wilcher B, Blankart CR, et al. Health Technology Assessment of Medical Devices: A Survey of Non-European Union Agencies. Int J Technol Assess Health Care 2015;31:154–65.

11. Garfield S, Polisena J, Spinner DS, Postulka A, Lu CY, Tiwana SK, et al. Health Technology Assessment for Molecular Diagnostics: Practices, Challenges, and Recommendations from the Medical Devices and Diagnostics Special Interest Group. Value Health 2016;19:577–87.

12. Bossuyt PM, Irwig L, Craig J, Glasziou P. Comparative Accuracy: Assessing New Tests against Existing Diagnostic Pathways. BMJ 2006;332:1089–92.

13. Lijmer JG, Leeflang M, Bossuyt PM. Proposals for a Phased Evaluation of Medical Tests. Med Decis Making 2009;29:E13–21.

14. National Institute of Health and Care Excellence. Diagnostics Assessment Programme Manual. Nice; December 2011. Available from: https://www.nice.org.uk/Media/Default/About/what-we-do/NICE-guidance/NICE-diagnostics-guidance/Diagnostics-assessment-programme-manual.pdf Last accessed 15th April 2022

15. Agency for Healthcare Research and Quality. Methods Guide for Medical Test Reviews. Ahrq Publication No. 12-Ec017. Rockville, Md: Agency for Healthcare Research and Quality; June 2012. Available from: https://effectivehealthcare.ahrq.gov/products/collections/methods-guidance-tests Last accessed 15th April 2022.

16. Oortwijn W, Jansen M, Baltussen R. Use of Evidence-Informed Deliberative Processes by Health Technology Assessment Agencies around the Globe. Int J Health Policy Manag 2020;9:27–33.

17. Shinkins B, Yang Y, Abel L, Fanshawe TR. Evidence Synthesis to Inform Model-Based Cost-Effectiveness Evaluations of Diagnostic Tests: A Methodological Review of Health Technology Assessments. BMC Med Res Methodol 2017;17:56.

18. Novielli N, Cooper NJ, Abrams KR, Sutton AJ. How Is Evidence on Test Performance Synthesized for Economic Decision Models of Diagnostic Tests? A Systematic Appraisal of Health Technology Assessments in the UK since 1997. Value Health 2010;13:952–7.

19. Grigore B, Ciani O, Dams F, Federici C, de Groot S, Möllenkamp M, et al. Surrogate Endpoints in Health Technology Assessment: An International Review of Methodological Guidelines. Pharmacoeconomics 2020;38:1055–70.

20. World Health Organisation. Institutionalization of Health Technology Assessment. Report on a Who Meeting; Bonn: Who Regional Office for Europe. 2001. https://www.euro.who.int/__data/assets/pdf_file/0016/120247/E72364.pdf Last accessed 15th April 2022.

21. GRADE. The Grade Working Group. Available from https://www.gradeworkinggroup.org/ Last accessed 15th April 2022.

22. WHO. World Health Organization. Available from https://www.who.int/ Last accessed 15th April 2022.

23. EUnetHTA. EUnetHTA Joint Action 2, Work Package 8. Hta Core Model ® Version 3.0 (Pdf); 2016. Available from http://www.htacoremodel.info/BrowseModel.aspx Last accessed 15th April 2022.

24. EUnetHTA. About EUnetHTA. Available from https://www.eunethta.eu/about-eunethta/ Last Accessed 15th April 2022.

25. Canadian Agency for Drugs and Technologies in Health. About the Health Technology Assessment Service, CADTH website: https://www.cadth.ca/about-cadth/what-we-do/products-services/hta (Last Updated 15th June 2021) Last accessed 15th April 2022.

26. Institute for Quality and Efficiency in Health Care. General methods (Version 6.0). Available from: https://www.iqwig.de/en/about-us/methods/methods-paper/ Last accessed 15th April 2022.

27. Medical Services Advisory Committee. Guidelines for preparing assessments for the Medical Services Advisory Committee. [Online]. Version 1.0. Available from: http://www.msac.gov.au/internet/msac/publishing.nsf/Content/Documents-for-Applicants-and-Assessment-Groups Last accessed 15th April 2022.

28. Statens beredning för medicinsk och social utärdering (Swedish Agency for Health Technology Assessment and assessment of social services). SBU Method Book (google translation into English), October 2020 [downloaded 10th February 2021]. Available from: https://www.sbu.se/en/method/ Last accessed 15th April 2022.

29. Zorginstituut Nederland. Medical tests (assessment of established medical science and medical practice). Report no 293. Zorginstituut Nederland: Diemen, 2011. Available from: https://english.zorginstituutnederland.nl/publications/reports/2011/01/20/medical-tests-assessment-of-established-medical-science-and-medical-practice Last accessed 15th April 2022.

30. U.S. Preventive Services Task Force. U.S. Preventive Services Task Force Procedure Manual. 2015. Available from: https://www.uspreventiveservicestaskforce.org/uspstf/about-uspstf/methods-and-processes/procedure-manual Last accessed 15th April 2022.

31. Medical Services Advisory Committee. Technical Guidelines for Preparing Assessment Reports for the Medical Services Advisory Committee – Service Type: Investigative (Version 3.0). July 2017. Canberra: Msac.

32. Heintz E, Lintamo L, Hultcrantz M, Jacobson S, Levi R, Munthe C, et al. Framework for Systematic Identification of Ethical Aspects of Healthcare Technologies: The SBU Approach. Int J Technol Assess Health Care 2015;31(3):124–130.

33. Takwoingi Y, Partlett C, Riley RD, Hyde C, Deeks JJ. Methods and Reporting of Systematic Reviews of Comparative Accuracy Were Deficient: A Methodological Survey and Proposed Guidance. J Clin Epidemiol 2020;121:1–14.

34. Yang B, Mallett S, Takwoingi Y, et al. Quadas-C: A Tool for Assessing Risk of Bias in Comparative Diagnostic Accuracy Studies. Ann Intern Med 2021;174:1592–9.

35. Ferrante di Ruffano L, Dinnes J, Sitch AJ, Hyde C, Deeks JJ. Test-Treatment RCTs Are Susceptible to Bias: A Review of the Methodological Quality of Randomized Trials That Evaluate Diagnostic Tests. BMC Med Res Methodol 2017;17:35.

